# Differential cognitive effects of unilateral left and right subthalamic nucleus deep brain stimulation for Parkinson disease

**DOI:** 10.1101/2023.02.27.23286478

**Authors:** Victor A Del Bene, Roy C. Martin, Sarah A. Brinkerhoff, Joseph W. Olson, Matthew J. Nelson, Dario Marotta, Christopher L. Gonzalez, Kelly A. Mills, Vidyulata Kamath, J. Nicole Bentley, Barton L. Guthrie, Robert T. Knight, Harrison C. Walker

## Abstract

**Objective:** To investigate hemispheric effects of directional versus ring subthalamic nucleus (STN) deep brain stimulation (DBS) surgery on cognitive function in patients with advanced Parkinson’s disease (PD).

**Methods:** We examined 31 PD patients (Left STN n = 17; Right STN n = 14) who underwent unilateral subthalamic nucleus (STN) DBS as part of a NIH-sponsored randomized, cross-over, double-blind (ring vs directional) clinical trial. Outcome measures were tests of verbal fluency, auditory-verbal memory, and response inhibition. First, all participants were pooled together to study the effects of directional versus ring stimulation. Then, we stratified the groups by surgery hemisphere and studied the longitudinal changes in cognition post-unilateral STN DBS.

**Results:** Relative to pre-DBS cognitive baseline performances, there were no group changes in cognition following unilateral DBS for either directional or ring stimulation. However, assessment of unilateral DBS by hemisphere revealed a different pattern. The left STN DBS group had lower verbal fluency than the right STN group (*t*(20.66 = -2.50, *p* = 0.02). Over a period of eight months post-DBS, verbal fluency declined in the left STN DBS group (*p* = 0.013) and improved in the right STN DBS group over time (*p* < .001). Similarly, response inhibition improved following right STN DBS (*p* = 0.031). Immediate recall did not significantly differ over time, nor was it affected by implant hemisphere, but delayed recall equivalently declined over time for both left and right STN DBS groups (left STN DBS *p* = 0.001, right STN DBS differ from left STN DBS *p* = 0.794).

**Conclusions:** Directional and ring DBS did not differentially or adversely affect cognition over time. Regarding hemisphere effects, verbal fluency decline was observed in those who received left STN DBS, along with the left and right STN DBS declines in delayed memory. The left STN DBS verbal fluency decrement is consistent with prior bilateral DBS research, likely reflecting disruption of the basal-ganglia-thalamocortical network connecting STN and inferior frontal gyrus. Interestingly, we found an improvement in verbal fluency and response inhibition following right STN DBS. It is possible that unilateral STN DBS, particularly in the right hemisphere, may mitigate cognitive decline.

## INTRODUCTION

Non-motor cognitive symptoms cause overwhelming disability in patients with advanced Parkinson’s disease (PD) ^1,2^. Ample evidence links PD with cognitive dysfunction, including declines in verbal fluency ^3,4^, and PD is associated with increasing risk of mild cognitive impairment and dementia over 5- and 10-year spans ^5,6^. Cognitive changes following deep brain stimulation (DBS) surgery for PD versus best medical therapy generally describe acceptable safety and tolerability ^7,8^. DBS at each of the major targets for movements disorders – subthalamic nucleus (STN), globus pallidus interna (GPi), and ventral intermediate thalamus (VIM) – improves motor symptoms but is associated with declines in semantic and phonemic verbal fluency ^9,10^. To date, most of the studies on cognitive outcomes after DBS focus on changes after simultaneous or immediately staged bilateral surgeries with conventional ring-shaped electrodes at the STN target. Whether directional leads and/or implant hemisphere impact cognitive function is unclear.

DBS does not appear to increase dementia risk ^11^, but bilateral surgeries for PD are associated with measurable declines in phonemic and semantic verbal fluency versus best medical therapy ^8,10^. Although ‘mild’ or ‘moderate’ from the standpoint of psychometrics, fluent verbal communication is integral to negotiating life activities, and DBS patients and their caregivers often complain about these functional declines when they occur. Changes in other domains of cognitive function following DBS are relatively understudied. Clear or consistent patterns of memory decline have thus far been elusive following STN DBS for PD ^7,8,12^, although STN stimulation may adversely impact behavioral inhibition ^13,14^. Further studies capturing changes in multidimensional cognition are therefore warranted in DBS patients, as the results could inform risk stratification in patients considering surgery and provide potential strategies to mitigate against functional declines. Furthermore, the impact of technological advances such as directional lead designs and potential closed-loop stimulation paradigms on cognitive function remains unclear ^15^.

Risk factors for cognitive declines after DBS surgery are incompletely understood ^7^. Prior studies are often limited by low statistical power, inconsistent outcome measures, and variable inclusion/exclusion criteria ^7,16^. Likely contributors include baseline cognitive performance, age, and the potential for stereotaxic lesion effects (independent from direct effects of stimulation itself) ^17-19^. Staged bilateral surgeries appear to be associated with verbal fluency changes, regardless of which brain hemisphere was targeted initially ^20^.

Language is perhaps the most well-recognized example of hemispheric brain lateralization^21^ and the motor manifestations of PD are often asymmetric, both at symptom onset and over time. Surprisingly, little is known about whether implant hemisphere impacts verbal fluency and other domains of cognitive function ^22,23^. SUNDIAL is a randomized, double-blind, cross-over study examining the safety and efficacy of unilateral directional versus ring STN DBS for PD. We first contrasted within-participant changes in multidomain cognitive function with novel directional stimulation versus conventional ring DBS. We then stratified patients by implant hemisphere, hypothesizing that left versus right STN DBS might differentially impact verbal fluency and other cognitive functions after surgery.

## METHODS

### Participants

We examined 31 PD patients (Table 1) who underwent unilateral STN DBS as part of a NIH-sponsored randomized, cross-over, double-blind clinical study (National Institute of Health BRAIN Initiative, clinicaltrials.gov NCT03353688). The United States Food and Drug Administration and The University of Alabama at Birmingham Institutional Review Board gave ethical approval for this work. All participants provided written informed consent prior to participation, only after a multidisciplinary committee recommended unilateral DBS at the STN target as part of routine care. Inclusion required >30% Movement Disorders Society – Unified Parkinson Disease Rating Scale (MDS-UPRDS) part 3 improvement after self-administration of dopaminergic medications versus the “off” state (>12 hours off dopaminergic medication) during a pre-operative screening visit. Other inclusion criteria included ages 18 - 70 years old, Hoehn and Yahr classification >1, and a Dementia Rating Scale-2 score ≥130 (out of 144). Exclusion criteria included duration of PD <4 years, history of stroke or other significant neurological conditions, and diagnosis of a functional movement disorder based on consensus criteria. Three screen failures did not experience >30% improvement in UPDRS part 3 motor score “off” versus on medications, one scored >25 on the Beck Depression Inventory (BDI-II), and the multidisciplinary DBS committee eventually recommended the GPi rather than the STN target in another. One of 31 enrollees (right STN case) underwent an uncomplicated surgery and later voluntarily withdrew from the study.

**TABLE 1:** Participant demographic factors.

|  |  | left STN |  |  | right STN |  |  | *p* |
| --- | --- | --- | --- | --- | --- | --- | --- | --- |
|  |  | sample (*n*) | mean | SD | sample (*n*) | mean | SD |  |
|  | age (years) | 17 | 56.7 | 8.6 | 14 | 58.9 | 6.3 | 0.145 |
|  | age at disease onset (years) | 17 | 49.3 | 8.6 | 14 | 50.8 | 6.7 | 0.593 |
|  | education (years) | 17 | 16.1 | 2.6 | 14 | 15.2 | 2.3 | 0.309 |
|  | Dementia Rating Scale | 16 | 138.4 | 3.5 | 11 | 138.5 | 2.7 | 0.989 |
|  | Beck Depression Inventory | 17 | 7.5 | 5.1 | 13 | 9.3 | 6.3 | 0.399 |
|  | Beck Anxiety Inventory | 15 | 10.5 | 6.5 | 8 | 13 | 4.8 | 0.313 |
|  | UPDRS motor off meds | 17 | 51.5 | 11.8 | 14 | 46.2 | 14.4 | 0.282 |
|  | UPDRS motor on meds | 17 | 25.6 | 7.5 | 13 | 25.9 | 9 | 0.915 |
|  | Hoehn & Yahr off meds | 17 | 2.1 | 0.3 | 14 | 2.1 | 0.4 | 0.843 |
|  | Hoehn & Yahr on meds | 17 | 2 | 0 | 13 | 2 | 0 | — |
|  | LEDD (mg) | 17 | 1115.8 | 508.8 | 14 | 776.9 | 338.5 | **0.035** |
|  | baseline to surgery (days) | 17 | 77.8 | 78.8 | 14 | 56.1 | 42.5 | 0.338 |
| handedness | right | 15 |  |  | 11 |  |  |  |
|  | left | 1 |  |  | 2 |  |  |  |
|  | ambidextrous | 1 |  |  | 1 |  |  | 0.717 |
| sex | male | 13 |  |  | 9 |  |  |  |
|  | female | 4 |  |  | 5 |  |  | 0.729 |
| race | white | 14 |  |  | 14 |  |  |  |
|  | black | 1 |  |  | 0 |  |  |  |
|  | asian | 2 |  |  | 0 |  |  | 0.255 |
| ethnicity | not hispanic or latino | 17 |  |  | 14 |  |  | — |

### Surgical procedure and stimulation parameters

Motor symptom asymmetry is a defining clinical feature of PD. Our routine clinical practice is to treat the most severely affected hemibody with unilateral DBS, followed by staged surgery on the opposite side of the brain (when, and if, needed) ^24-28^. The same neurosurgeons (BG, JNB) implanted a 1-3-3-1 directional lead (Boston Scientific Vercise DBS system, Natick MA, USA, FDA IDE# G170063) at the STN target under local anesthesia with the patient fully awake. Midazolam 1-2 mg is administered during the placement of the stereotactic frame. Pre-surgical brain MRI scans are co-registered with intraoperative O-arm CT images for STN targeting and to assess micro- and macroelectrode locations. The final DBS location is based on both awake electrophysiology recordings and the co-registered MRI and CT anatomic images. On average, there were 35.2 ± 21.5 days between lead implant and device activation. Participants were allocated to directional or ring stimulation at 2- and 4-month follow-up intervals in a double-blind fashion using block randomization in RedCap using an embedded randomization process. Regardless of directional or ring stimulation, programming goals were to maximize improvements in PD motor symptoms.

### Neuropsychological assessments

All participants completed a standardized, comprehensive neuropsychological battery suitable for patients considering DBS ^25^. Baseline screening assessments on medications were discussed at a multidisciplinary DBS consensus conference prior to recruitment and enrollment. COVID-19 emerged mid-trial, such that many encounters were converted to a telehealth format over a HIPAA-compliant two-way video connection (Zoom Communications), consistent with standard clinical practices during the pandemic ^29,30^. Whether in person or via telehealth, participants were in a quiet, distraction-free environment, and writing or note-taking was not allowed. Remote assessments utilized computer displays with sufficient screen area to allow easy viewing of DKEFS Color-Word Inhibition Test stimuli. All other tests did not require visual stimulus presentation.

We examined the following cognitive outcomes: *Dementia Screen*. Dementia Rating Scale (DRS-2) ^31^ served as a general cognitive screen at the pre-operative baseline encounter only. *Phonemic Verbal Fluency*. The F-A-S version was administered at the pre-surgery baseline assessment, and subsequent study visits alternated between the C-F-L and F-A-S versions ^32^. Participants were asked to generate as many words as possible starting with a given letter over a 60-second period. *Immediate and Delayed Memory*. The Rey Auditory Verbal Learning Test (RAVLT) ^33^ is a 15-item word-list learning test. Outcomes were learning trials (1-5) total score and long-delayed recall number of words. Pre-surgery sessions used form AB, and visits 2, 3, and 4 used versions CB, Cr-AB, and Ge-AB, respectively. *Response Inhibition*. The Delis-Kaplan Executive Function System (DKEFS) ^34^ Color-Word Interference Test Trial 3 (Inhibition) evaluates response inhibition. Color naming, word reading, inhibition, and inhibition/switching were examined at each encounter, and the primary outcome for this study was time-to-completion for the inhibition trial in seconds. For all neuropsychological tests, raw scores were used for the primary analyses.

### Statistical analyses

R was used for all analyses (R Core Team, 2020; R Studio Team, 2021). Baseline continuous and categorical demographic variables were compared by implant hemisphere using Welch’s two-sample t-tests and chi-square tests (**Table 1**). To evaluate baseline cognition by implant hemisphere, Welch’s two-sample t-tests compared baseline cognitive function by implant hemisphere. To evaluate within-participant changes in cognitive function by stimulation, binomial tests determined the probability that directional stimulation resulted in better cognitive performance than ring stimulation. To evaluate the effect of DBS on cognitive function over time, linear mixed effects regression (LMER) models ^35^ estimated longitudinal change in cognitive function regardless of implant hemisphere. To determine if DBS hemisphere differentially impacts cognitive function after surgery, LMER models estimated longitudinal change in cognitive function by implant hemisphere, for both the raw cognitive scores and the percent change from baseline. Raw score models included a random intercept by participant and percent change models included a random slope by participant. All models included covariates for education level (years) and interaction terms for hemisphere by time. Percent change models also included a slope covariate for baseline score. All models for the DKEFS Color-Word Inhibition Trial also included covariates for color naming speed and word naming speed. Visit type (telehealth versus in-person) and daily levodopa equivalent dose (LEDD) were tested as covariates to each model, but neither significantly improved model fit to the data for any outcome; therefore, visit type and LEDD were not considered in the models. Fixed effects were considered significant *a priori* with *p*-values of <0.05.

## RESULTS

### Baseline demographics and cognitive performance

Age, age at disease onset, duration of disease, education, DRS-2 total, Beck Depression Inventory total, Beck Anxiety Index total, MDS-UPDRS part 3 score on and “off” medications, days from baseline screening to surgery, handedness, sex, race, and ethnicity did not differ by implant hemisphere (**Table 1**). Participants with predominant motor symptoms on the right body who received left STN DBS had higher LEDD at baseline. Participants with predominant symptoms on the left body who received right STN DBS displayed better preoperative verbal fluency function than those with left implants (*t*(20.66 = -2.50, *p* = 0.02). Immediate recall (*t*(22.74 = -0.57, *p* = 0.57), delayed recall (*t*(28.97 = 1.25, *p* = 0.22), and response inhibition (*t*(27.98 = 0.67, *p* = 0.51) did not differ by implant hemisphere at baseline. On average, the final cognitive assessment occurred at 253 days or 8 months after DBS surgery.

### Cognitive performance following unilateral STN DBS in either hemisphere

When looking at the entire sample (pooling across hemispheres) and ignoring stimulation configuration (i.e., directional versus ring), unilateral STN DBS did not significantly impact verbal fluency, response inhibition, immediate recall, or delayed recall over an average follow-up duration of eight months after surgery (**Figure 1A, Table 2**). Inhibition processing speed was faster in patients who also performed DKEFS color naming (p = 0.008) and word naming (p = 0.006) faster.

**Table 2.** Statistical models of overall cognitive performance following unilateral STN DBS.

|  | **Verbal Fluency** | **Response Inhibition** | **Immediate Recall** | **Delayed Recall** |
| --- | --- | --- | --- | --- |
| Intercept | **43.351 (2.392)\*\*\*** | **30.839 (5.306)\*\*\*** | **47.879 (1.511)\*\*\*** | **9.788 (0.566)\*\*\*** |
| Education (years) | -0.387 (0.940) | -0.258 (0.747) | -0.220 (0.544) | -0.027 (0.203) |
| Color Naming Speed (sec) | — | **0.442 (0.164)\*\*** | — | — |
| Word Naming Speed (sec) | — | **0.599 (0.213)\*\*** | — | — |
| Change over time (days) | -0.003 (0.006) | -0.003 (0.006) | -0.004 (0.006) | -0.004 (0.002) |
| Num. obs. | 115 | 109 | 114 | 111 |
| Num. groups: ID | 31 | 30 | 31 | 30 |
| Var: ID (Intercept) | 149.299 | 86.613 | 43.640 | 6.175 |
| Var: Residual | 37.746 | 26.894 | 36.407 | 4.655 |

*Note: Estimates are given in Coefficient (SEM). The intercept indicates the score at baseline (0 days) for a patient with an average education level (15.7 years) (and, for response inhibition, a patient with an average color naming speed (31.6 sec) and word naming speed (22.2 sec)). Asterisks indicate significant difference from zero. \* $p < 0.05$ , \*\* $p < 0.01$ , \*\*\* $p < 0.001$ .*

**Figure 1.**
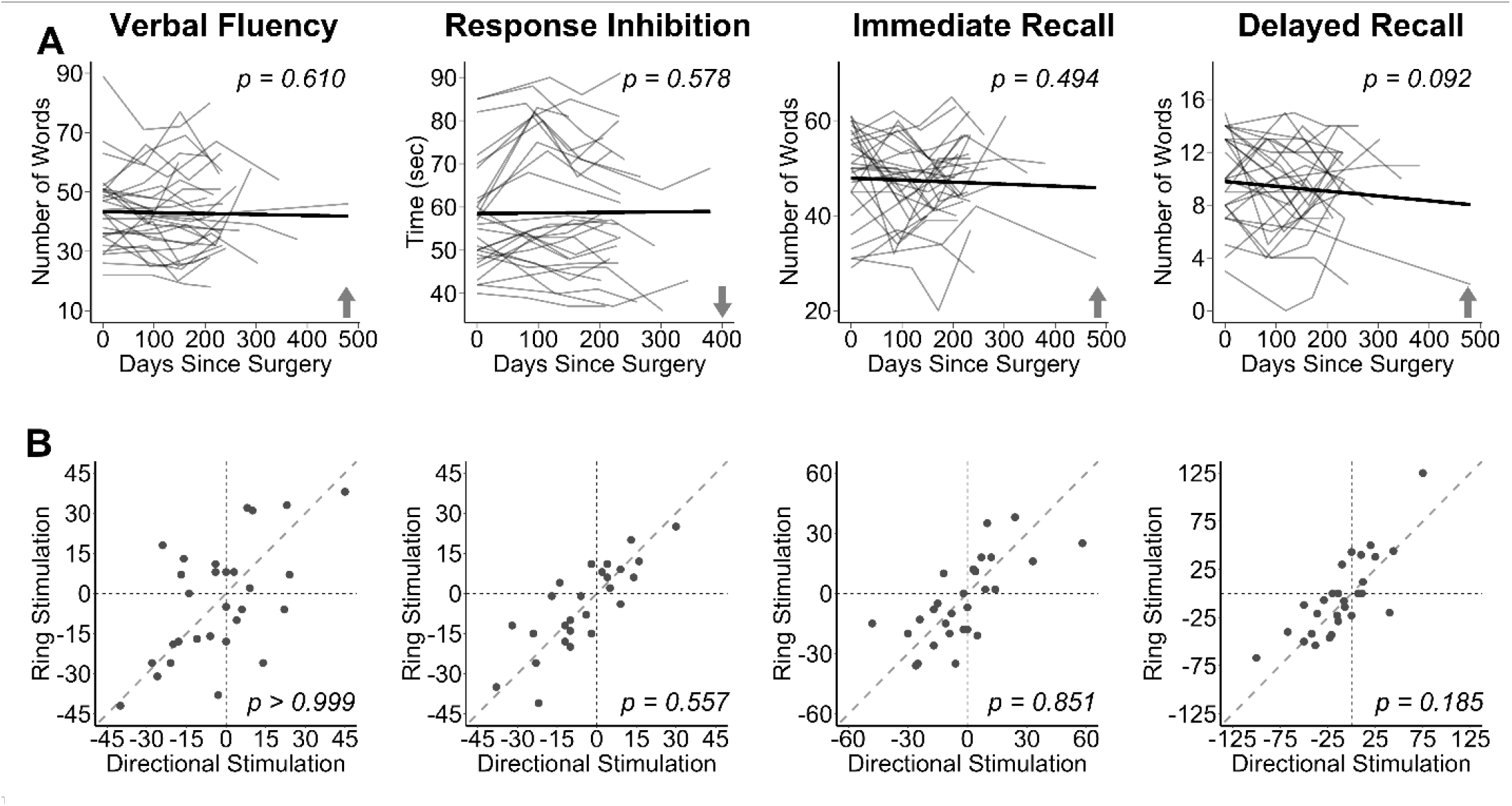
Cognitive performance by Stimulation. **(A)** Line segments show within-participant changes in cognitive performance over time after DBS surgery, and bold lines show sample means. On tests of verbal fluency and memory, higher scores reflect improvement, while a lower score on response inhibition reflects an improved (i.e., faster) performance. p-values indicate significance of slopes over time. **(B)** Unity plots contrast cognitive performance with double-blind comparison of ring versus directional stimulation. p-values indicate significance of the binomial test in improvement on either stimulation setting.

### Cognitive performance by stimulation mode

In double-blind assessments of directional versus ring DBS, stimulation configuration (directional versus ring) did not differentially modify verbal fluency, response inhibition, immediate recall, and delayed recall at our level of statistical power (**Figure 1B, Table 3**). Compared to ring stimulation, directional STN DBS was associated with verbal fluency improvements in 52% of the cases (*n* = 29, *p*>0.999); faster response inhibition in 42% of the cases (*n* = 26, *p*=0.557); greater immediate recall in 54% of the cases (*n* = 28, *p*=0.851); and greater delayed recall in 36% of the cases (*n* = 28, *p*=0.185).

**Table 3.** Binomial tests of cognitive performance by stimulation mode.

| ***n*** | **Verbal Fluency** | **Response Inhibition** | **Immediate Recall** | **Delayed Recall** |
| --- | --- | --- | --- | --- |
| Total sample | 29 | 26 | 28 | 28 |
| Better on directional | 15 | 11 | 15 | 10 |
| Better on ring | 13 | 12 | 13 | 13 |
| Same on directional and ring | 1 | 3 | 0 | 5 |
| *Exact Binomial Test* | 0.52 ($p>0.999$) | 0.42 ($p=0.557$) | 0.54 ($p=0.851$) | 0.36 ($p=0.185$) |

*Note: Exact binomial test statistic indicates the probability of directional stimulation yielding better performance. $p$ -values indicate the probability that the true probability of directional stimulation yielding better performance is not equal to 0.5.*

### Cognitive performance by implant hemisphere

The summary of results from mixed models are provided in **Table 4** (raw scores) and **Table 5** (percent change). Implant hemisphere significantly affected longitudinal verbal fluency function. Following unilateral right STN DBS, verbal fluency improved over time with an increase of 1% of baseline score every 14.7 days (*p* < 0.001), or 1 word per 75.4 days after surgery (*p* = 0.021). Following unilateral left STN DBS, verbal fluency worsened over time with a decrease of 1% of baseline score every 19.0 days (*p* = 0.01) (**Figure 2**).

**Table 4.**
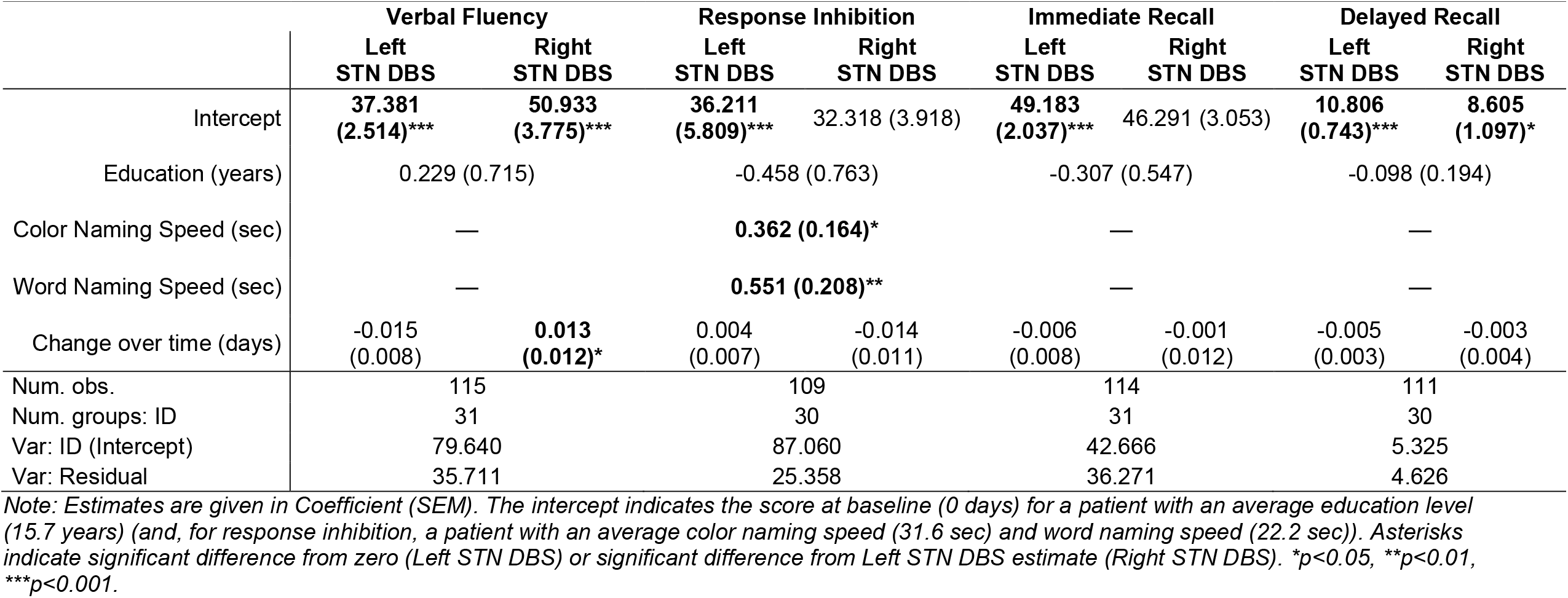
Statistical models of raw cognitive performance by implant hemisphere.

|  | Verbal Fluency |  | Response Inhibition |  | Immediate Recall |  | Delayed Recall |  |
| --- | --- | --- | --- | --- | --- | --- | --- | --- |
|  | Left STN DBS | Right STN DBS | Left STN DBS | Right STN DBS | Left STN DBS | Right STN DBS | Left STN DBS | Right STN DBS |
| Intercept | **37.381**<br>(2.514)*** | **50.933**<br>(3.775)*** | **36.211**<br>(5.809)*** | 32.318 (3.918) | **49.183**<br>(2.037)*** | 46.291 (3.053) | **10.806**<br>(0.743)*** | **8.605**<br>(1.097)* |
| Education (years) | 0.229 (0.715) |  | -0.458 (0.763) |  | -0.307 (0.547) |  | -0.098 (0.194) |  |
| Color Naming Speed (sec) | — |  | **0.362 (0.164)\*** |  | — |  | — |  |
| Word Naming Speed (sec) | — |  | **0.551 (0.208)\*\*** |  | — |  | — |  |
| Change over time (days) | -0.015<br>(0.008) | **0.013**<br>(0.012)* | 0.004<br>(0.007) | -0.014<br>(0.011) | -0.006<br>(0.008) | -0.001<br>(0.012) | -0.005<br>(0.003) | -0.003<br>(0.004) |
| Num. obs. | 115 |  | 109 |  | 114 |  | 111 |  |
| Num. groups: ID | 31 |  | 30 |  | 31 |  | 30 |  |
| Var: ID (Intercept) | 79.640 |  | 87.060 |  | 42.666 |  | 5.325 |  |
| Var: Residual | 35.711 |  | 25.358 |  | 36.271 |  | 4.626 |  |

*Note: Estimates are given in Coefficient (SEM). The intercept indicates the score at baseline (0 days) for a patient with an average education level (15.7 years) (and, for response inhibition, a patient with an average color naming speed (31.6 sec) and word naming speed (22.2 sec)). Asterisks indicate significant difference from zero (Left STN DBS) or significant difference from Left STN DBS estimate (Right STN DBS). \* $p < 0.05$ , \*\* $p < 0.01$ , \*\*\* $p < 0.001$ .*

**Table 5.** Statical models of percent change cognitive performance by implant hemisphere.

|  | Verbal Fluency |  | Response Inhibition |  | Immediate Recall |  | Delayed Recall |  |
| --- | --- | --- | --- | --- | --- | --- | --- | --- |
|  | Left<br>STN DBS | Right<br>STN DBS | Left<br>STN DBS | Right<br>STN DBS | Left<br>STN DBS | Right<br>STN DBS | Left<br>STN DBS | Right<br>STN DBS |
| Intercept | -3.089<br>(2.536) | 4.392<br>(3.809) | 5.897<br>(6.298) | 3.514<br>(2.464) | -0.742<br>(2.341) | -0.216<br>(3.509) | -4.235<br>(5.732) | 3.649<br>(8.470) |
| Education (years) | 0.087 (0.589) |  | 0.126 (0.455) |  | -0.407 (0.590) |  | -0.388 (1.349) |  |
| Color Naming Speed<br>(sec) | — |  | -0.079 (0.197) |  | — |  | — |  |
| Word Naming Speed<br>(sec) | — |  | -0.326 (0.273) |  | — |  | — |  |
| Change over time (days) | **-0.053**<br>**(0.020)\*\*** | **0.068**<br>**(0.033)\*\*\*** | -0.052<br>(0.026) | **0.012**<br>**(0.029)\*** | 0.031<br>(0.024) | 0.039<br>(0.033) | **-1.040**<br>**(0.282)\*\*\*** | -1.056<br>(0.074) |
| Days * Baseline value | **-0.004 (0.001)\*\*\*** |  | 0.001 (0.001) |  | **-0.006 (0.001)\*\*\*** |  | **-0.034 (0.009)\*\*\*** |  |
| Num. obs. | 115 |  | 109 |  | 114 |  | 111 |  |
| Num. groups: ID | 31 |  | 30 |  | 31 |  | 30 |  |
| Var: ID (days) | 0.002 |  | 0.004 |  | 0.004 |  | 0.015 |  |
| Var: Residual | 143.961 |  | 52.246 |  | 119.856 |  | 688.807 |  |

*Note: Estimates are given in Coefficient (SEM). The intercept indicates the score at baseline (0 days) for a patient with an average education level (15.7 years) and an average baseline score (and, for response inhibition, a patient with an average color naming speed (31.6 sec) and word naming speed (22.2 sec)). Asterisks indicate significant difference from zero (Left STN DBS) or significant difference from Left STN DBS estimate (Right STN DBS). \* $p < 0.05$ , \*\* $p < 0.01$ , \*\*\* $p < 0.001$ .*

**Figure 2.**
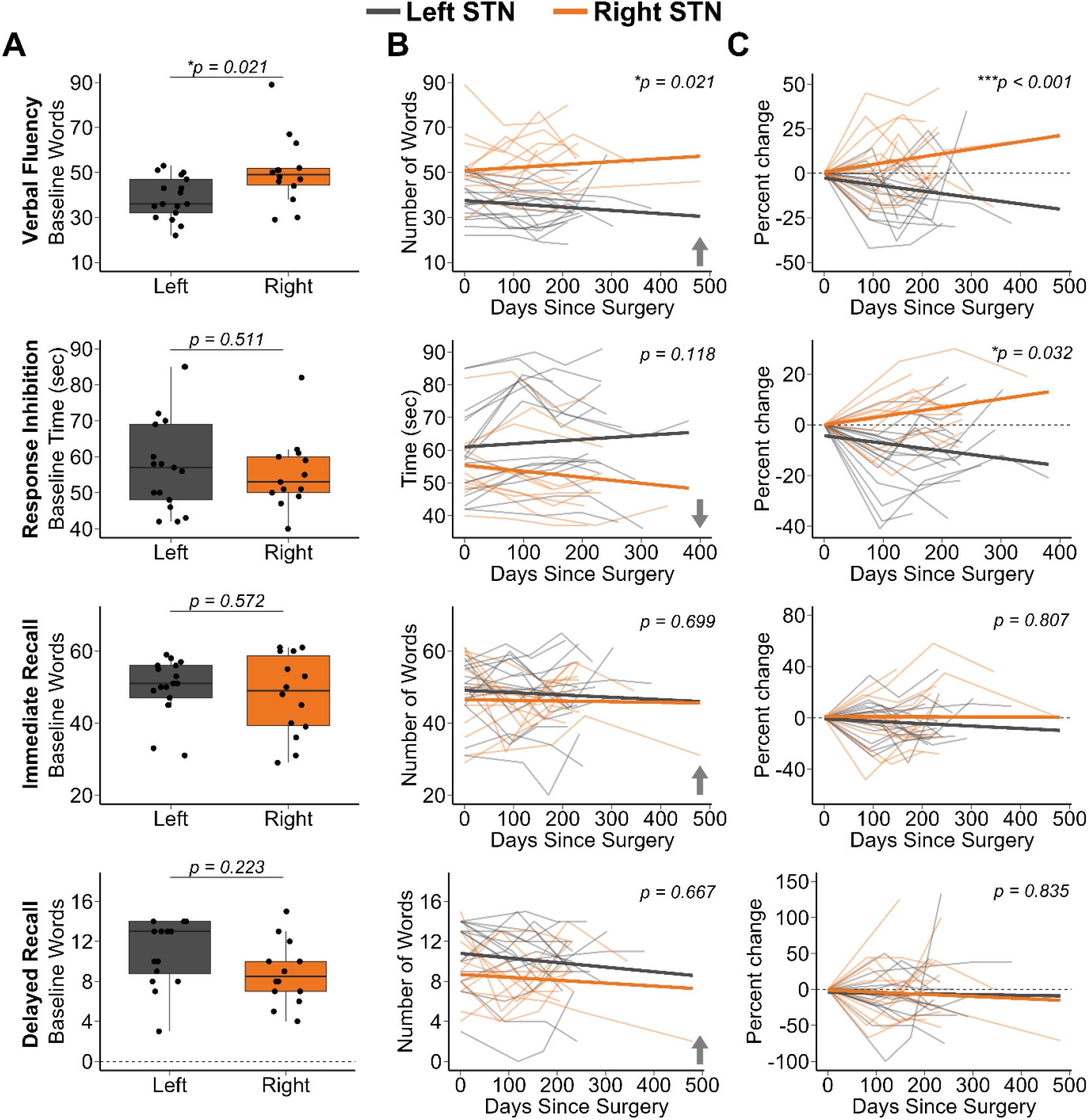
Baseline and longitudinal cognitive function in unilateral STN DBS patients. **(A)** Boxplots of baseline multi-domain cognitive performance by implant hemisphere. p-values indicate significance between groups at baseline. **(B, C)** Line segments show within-participant changes in cognitive performance over time by implant hemisphere, and bold lines show group means. On raw scores of tests of verbal fluency and memory, higher scores reflect improvement, while a lower score on a test of inhibition reflects an improved (i.e., faster) performance. On percent change of all tests, a positive change reflects improvement while a negative change reflects worsening. *p*-values indicate significant difference between groups in slopes over time.

Response inhibition improved following right unilateral STN DBS by 1% of baseline score every 83 days (*p* = 0.032) (**Figure 2**). Regardless of implant hemisphere, raw response inhibition processing speed was faster in patients who also performed DKEFS color naming (p = 0.029) and word naming (p = 0.010) faster. Immediate recall did not significantly differ over time, nor was it affected by implant hemisphere (**Figure 2**). Delayed recall worsened following both left and right unilateral STN DBS by 1% of baseline score every 1 day (left STN DBS *p* = 0.001, right STN DBS differ from left STN DBS *p* = 0.835).

### Functional impairments in verbal fluency over time

We evaluated the rates of clinical verbal fluency impairment by implant hemisphere at baseline and over the eight months following unilateral STN DBS (**Table 6**). Here, impairment is defined as either 1 or 1.5 standard deviations below the 50^th^ percentile (i.e., the 16^th^ and 5^th^ percentiles, respectively) of a normative sample ^32^. A proportion of left hemisphere implants had baseline and longitudinal verbal fluency impairments, whereas right STN implants had little, if any, impairments. Statistically significant group level verbal fluency declines do not uniformly impair all patients who receive left STN DBS.

**TABLE 6:**
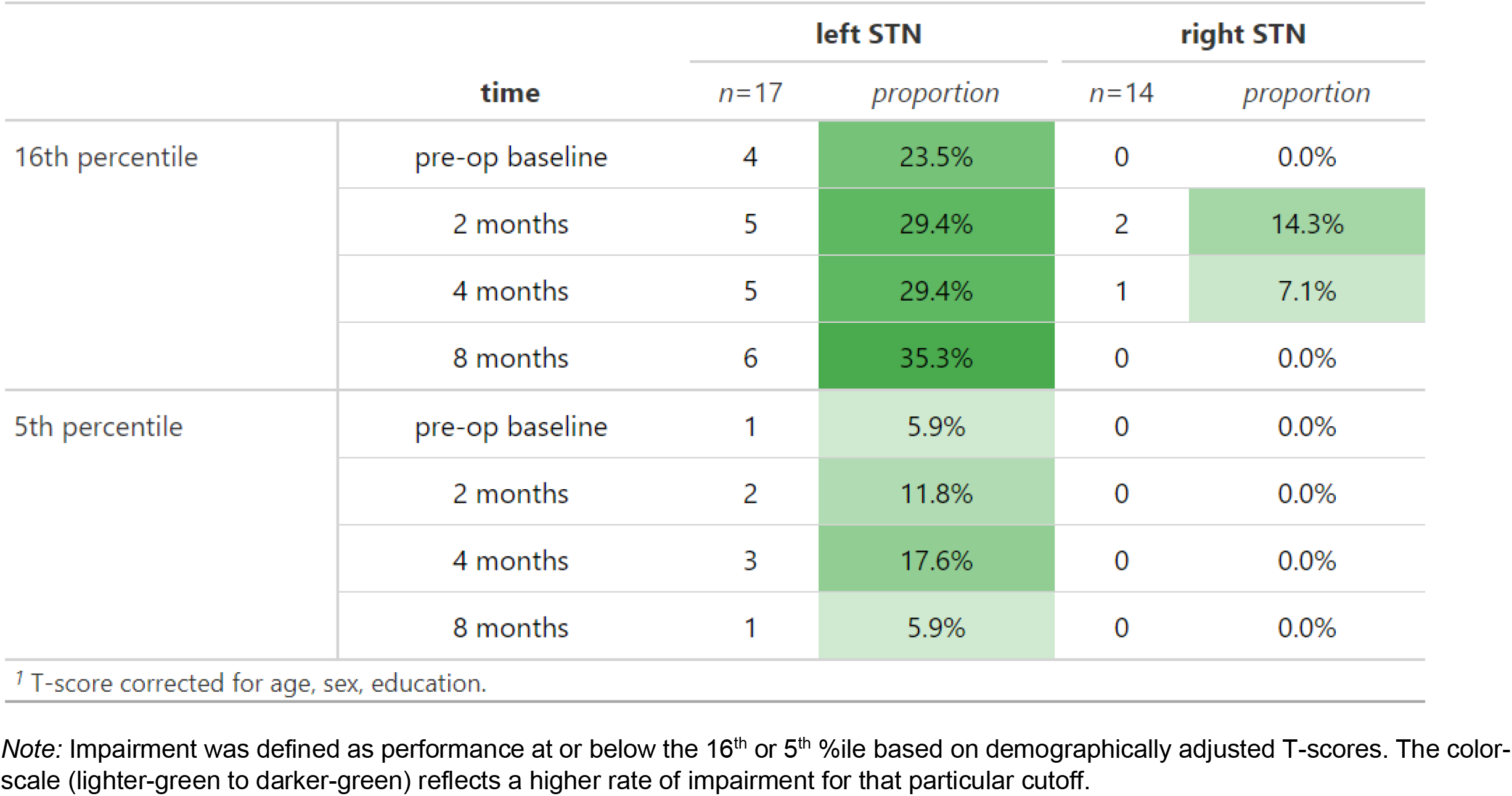
Verbal fluency impairment by implant hemisphere across clinical thresholds^*1*^.

## DISCUSSION

Declines in verbal fluency are the most commonly reported neuropsychological sequela of DBS surgery for movement disorders. While PD-related verbal fluency decline is also common due to disease progression ^36^ declines are typically more pronounced following DBS than those observed in best medical therapy groups ^10^. Across the entire PD sample in our study (left and right pooled together), double-blind, randomized crossover comparison of directional versus ring stimulation showed no significant changes across each cognitive domain. Changes in verbal fluency were more strongly driven by disease laterality and implant hemisphere, with worse baseline verbal fluency and greater declines in verbal fluency following unilateral STN DBS in the presumed language-dominant left hemisphere. In contrast, unilateral right STN implants were associated with better baseline verbal fluency and even group-level improvements at up to 8 months after surgery. Similarly, we found improved response inhibition speed over time following right STN DBS. When comparing left and right STN DBS cases, a modest decline in delayed auditory-verbal memory was also observed. Our findings raise the hypothesis that when medically appropriate, unilateral STN DBS, rather than bilateral DBS, could be a modifiable risk factor for preventing cognitive decline associated with DBS therapy for PD.

Non-motor effects of unilateral DBS are a relatively neglected research topic. Independent of implant hemisphere, cognition in PD patients was not affected significantly by unilateral STN DBS at the group level at our level of statistical power, either versus baseline or when contrasting ring versus directional stimulation paradigms. This contrasts with a substantial prior literature reporting measurable declines in verbal fluency declines following bilateral STN DBS versus best medical therapy ^10,12,37,38^. To the best of our knowledge, this is the first study characterizing cognitive performance in response to directional brain stimulation. Directional stimulation offers a wider therapeutic window than more traditional omnidirectional DBS ^39,40^, and directional stimulation is typically preferred by patients at the STN target ^40^. Our findings suggest that either ring or directional unilateral STN stimulation optimized for motor improvement likely has a favorable cognitive safety profile in patients with PD.

Hemispheric lateralization of motor symptoms may be associated with distinct cognitive phenotypes in PD patients with advanced-staged disease warranting consideration for DBS. For example, our left STN DBS sample at baseline had lower verbal fluency performance than our right STN DBS group. Our findings demonstrate that these potential cognitive phenotypes likely interact differentially with unilateral STN DBS. The magnitude of verbal fluency declines following unilateral left STN DBS in our sample are compatible with published declines following bilateral DBS ^10,41,42^, and with prior observations that have linked left STN lead location with changes in verbal fluency output ^41^. Collectively, these findings suggest that the left hemisphere lead likely drives most of the observed verbal fluency declines following bilateral surgeries. Our finding of relative improvements in longitudinal verbal fluency performance following right STN DBS is novel and somewhat surprising, although a prior study suggested similar hemispheric effects with a more limited sample ^23^. Interestingly, another study examined staged bilateral STN and GPi DBS and pallidotomy, and reported postoperative declines in verbal fluency following initial left but not right hemisphere targeting ^20^. Moreover, patients with initial right hemisphere targeting who went on to have left STN or GPi, experienced declines in verbal fluency performance following this second surgery ^20^. In the lesioning literature, unilateral pallidotomy and thalamotomy studies report greater declines in verbal fluency following left hemisphere procedures ^43,44^, but one pallidotomy study reported a non-significant trend towards improved verbal fluency following right hemisphere procedures with a smaller sample ^45^.

We found relatively faster response inhibition speed following right STN DBS, and like the improved verbal fluency post-right STN DBS, represents a novel finding. The published literature on post-DBS performance on the Stroop-Color Word Test and the DKEFS Color-Word Inhibition Test generally show a decline following bilateral stimulation of the STN ^46,47^. This improvement may indicate faster cognitive processing speed following unilateral right STN DBS, as evidenced by faster response inhibition performance and improved verbal fluency. Alternatively, this could also indicate the absence of post-DBS decline in the right STN cases along with practice effect.

In our sample, we found no evidence for hemispheric differences in immediate memory recall at our level of statistical power. In contrast, we found a modest statistically significant decline in auditory-verbal delayed recall for both left and right STN DBS cases. Compared to the prior literature, a meta-analysis of neuropsychological data from 26 STN DBS studies detected small declines in auditory-verbal learning and memory (average random effect size = 0.21), although the average random effect size for verbal fluency in the meta-analysis study was 0.64, reflecting a much stronger effect ^38^. A more recent study also reported mild declines in auditory-verbal memory, but their sample consisted of only bilateral STN cases ^48^. While we found a similar pattern of delayed memory decline following unilateral STN DBS, the effect size is smaller in our sample. One possibility is that bilateral DBS poses a marginally greater risk to memory function than unilateral surgery, but prospective, randomized studies are required to make definitive conclusions.

Disruption of the basal-ganglia-thalamocortical network likely underlies declines in verbal fluency following DBS, although the specific nature of the relevant interactions is unclear. Single unit studies provide evidence for functional roles of the STN in speech production ^49^, and both animal and human models have identified monosynaptic pathways from prefrontal cortex and temporal lobe to STN ^50,51^. Functional and structural neuroimaging studies consistently support a role for the left inferior gyrus during verbal fluency tasks ^52,53^, which has direct connections with the dorsal STN, as well ^51^. Additionally, evoked potential studies provide evidence for direct connections between STN and both inferior frontal gyrus and superior temporal gyrus and the opercular speech network ^50^. Outside of intrinsic STN function are considerations related to off-target stimulation effects or lesion effects from physical perturbation of cortical and subcortical elements in the lead trajectory prior to initiating therapy. Notably, microlesion effects contributed significantly to verbal fluency declines in one sham-controlled trial ^17^. Another study found an association between trajectory angle and verbal fluency declines. Specifically, a lateral point of entry through the left superior frontal gyrus was associated with higher levels of verbal fluency decline from baseline, with no change in verbal fluency observed when the approach is through the caudate nucleus ^54^, although others have shown that caudate penetration is associated with increased risk of decline in global cognition and working memory ^18^ It is likely that the microlesion effect in the left hemisphere disrupts the basal-ganglia-thalamocortical network, yielding verbal fluency declines, which may potentially be exacerbated by the stimulator depending on the DBS lead location ^41,55^. A similar mechanism may explain the mild declines in auditory-verbal memory retrieval seen in our sample ^56^.

Our study design has notable strengths and some important limitations. Although we are not aware of another study with as large a sample size comparing hemispheric effects of unilateral DBS on cognitive function, our sample size is still modest ^16^ and larger multicenter studies might detect additional or more subtle hemispheric effects on cognitive function. Also, we did not directly contrast unilateral DBS versus either best medical therapy or bilateral DBS; therefore, inferences across samples from independent studies should be interpreted with some caution. For example, since those receiving left STN DBS also had greater LEDD values reflecting greater severity of motor symptoms, our cognitive findings could be related to manifestations of PD severity itself, rather than neurostimulation effects, and we are unable to rule this out without a non-stimulated cohort. Regardless, we observed no significant group level changes in cognition in our unilateral sample when target hemisphere was not included in the statistical model, whereas most prior studies with reported significant group-level verbal fluency declines following bilateral DBS with smaller samples. Furthermore, we observed verbal fluency and response inhibition improvements following right STN DBS, which would not be expected in any case based on the prior literature. While these modest improvements might in part reflect practice effects on the task, at the very least, they do not represent declines post-DBS. Moreover, the left STN DBS group did not show practice effects on the verbal fluency task, but rather, showed decline, and we used alternate forms to mitigate this possibility. Finally, while our study team was double-blinded to the stimulation parameters, patient and examiner were not blinded to implant hemisphere. That said, changes in cognitive function were noted at multiple time points and were generally consistent within individuals.

We conclude that unilateral directional and ring stimulation at the STN target are equally safe and do not differentially and adversely affect cognition in patients with PD. Our examination of unilateral rather than bilateral DBS surgery allowed the identification of notable verbal fluency changes following left and not right hemisphere STN DBS surgery. Both implant hemisphere and the nature of PD itself likely impact verbal fluency over time. Similarly, we found right STN DBS improved response inhibition speed in our sample. A modest decline in delayed auditory-verbal memory was observed, consistent with the prior bilateral DBS studies. Future studies should prospectively contrast unilateral and bilateral DBS with best medical therapy to develop more personalized therapies to optimize motor and non-motor function in patients with PD and could also study the brain networks involved in verbal fluency decline associated with STN DBS, as previously done with DBS-associated working memory change ^57^.

## Data Availability

As part of the NIH BRAIN Initiative, all data are available online.

https://dabi.loni.usc.edu/data

## Notes

### Competing Interest Statement

The authors have declared no competing interest.

### Clinical Trial

NCT03353688

### Funding Statement

We are grateful for funding from the National Institutes of Health BRAIN Initiative (1UH3NS100553) and the Michael J. Fox Foundation (both to HW).

### Author Declarations

The United States Food and Drug Administration and The University of Alabama at Birmingham Institutional Review Board gave ethical approval for this work.

## References

1. Antonini A, Barone P, Marconi R, et al. The progression of non-motor symptoms in Parkinson’s disease and their contribution to motor disability and quality of life. Journal of neurology. 2012;259(12):2621–2631.

2. Bugalho P, Ladeira F, Barbosa R, et al. Progression in Parkinson’s Disease: Variation in Motor and Non-motor Symptoms Severity and Predictors of Decline in Cognition, Motor Function, Disability, and Health-Related Quality of Life as Assessed by Two Different Methods. Movement Disorders Clinical Practice. 2021;8(6):885–895.

3. Baiano C, Barone P, Trojano L, Santangelo G. Prevalence and clinical aspects of mild cognitive impairment in Parkinson’s disease: A meta-analysis. Movement Disorders. 2020;35(1):45–54.

4. Wallace ER, Segerstrom SC, van Horne CG, Schmitt FA, Koehl LM. Meta-analysis of cognition in Parkinson’s disease mild cognitive impairment and dementia progression. Neuropsychology Review. 2022;32(1):149–160.

5. Pedersen KF, Larsen JP, Tysnes O-B, Alves G. Natural course of mild cognitive impairment in Parkinson disease: A 5-year population-based study. Neurology. 2017;88(8):767–774.

6. Aarsland D, Kurz MW. The epidemiology of dementia associated with Parkinson disease. Journal of the neurological sciences. 2010;289(1-2):18–22.

7. Tröster AI. Some clinically useful information that neuropsychology provides patients, carepartners, neurologists, and neurosurgeons about deep brain stimulation for Parkinson’s disease. Archives of Clinical Neuropsychology. 2017;32(7):810–828.

8. Wang J, Pan R, Cui Y, Wang Z, Li Q. Effects of Deep Brain Stimulation in the Subthalamic Nucleus on Neurocognitive Function in Patients With Parkinson’s Disease Compared With Medical Therapy: A Meta-Analysis. Frontiers in Neurology. 2021;12:610840.

9. Lachenmayer ML, Mürset M, Antih N, et al. Subthalamic and pallidal deep brain stimulation for Parkinson’s disease—meta-analysis of outcomes. NPJ Parkinson’s disease. 2021;7(1):1–10.

10. Wyman-Chick KA. Verbal fluency in Parkinson’s patients with and without bilateral deep brain stimulation of the subthalamic nucleus: a meta-analysis. Journal of the International Neuropsychological Society. 2016;22(4):478–485.

11. Bove F, Fraix V, Cavallieri F, et al. Dementia and subthalamic deep brain stimulation in Parkinson disease: A long-term overview. Neurology. 2020;95(4):e384–e392.

12. Woods SP, Fields JA, Tröster AI. Neuropsychological sequelae of subthalamic nucleus deep brain stimulation in Parkinson’s disease: a critical review. Neuropsychology review. 2002;12(2):111–126.

13. Witt K, Pulkowski U, Herzog J, et al. Deep brain stimulation of the subthalamic nucleus improves cognitive flexibility but impairs response inhibition in Parkinson disease. Archives of neurology. 2004;61(5):697–700.

14. Bucur M, Papagno C. Deep Brain Stimulation in Parkinson Disease: A Meta-analysis of the Long-term Neuropsychological Outcomes. Neuropsychology Review. 2022:1–40.

15. Krauss JK, Lipsman N, Aziz T, et al. Technology of deep brain stimulation: current status and future directions. Nature Reviews Neurology. 2021;17(2):75–87.

16. Woods SP, Rippeth JD, Conover E, Carey CL, Parsons TD, Tröster AI. Statistical power of studies examining the cognitive effects of subthalamic nucleus deep brain stimulation in Parkinson’s disease. The Clinical Neuropsychologist. 2006;20(1):27–38.

17. Lefaucheur R, Derrey S, Martinaud O, et al. Early verbal fluency decline after STN implantation: is it a cognitive microlesion effect? Journal of the neurological sciences. 2012;321(1-2):96–99.

18. Witt K, Granert O, Daniels C, et al. Relation of lead trajectory and electrode position to neuropsychological outcomes of subthalamic neurostimulation in Parkinson’s disease: results from a randomized trial. Brain. 2013;136(7):2109–2119.

19. Costentin G, Derrey S, Gérardin E, et al. White matter tracts lesions and decline of verbal fluency after deep brain stimulation in Parkinson’s disease. Human Brain Mapping. 2019;40(9):2561–2570.

20. Rothlind JC, Cockshott RW, Starr PA, Marks WJ. Neuropsychological performance following staged bilateral pallidal or subthalamic nucleus deep brain stimulation for Parkinson’s disease. Journal of the International Neuropsychological Society. 2007;13(1):68–79.

21. Riès SK, Dronkers NF, Knight RT. Choosing words: left hemisphere, right hemisphere, or both? Perspective on the lateralization of word retrieval. Annals of the New York Academy of Sciences. 2016;1369(1):111–131.

22. Alberts JL, Hass CJ, Vitek JL, Okun MS. Are two leads always better than one: an emerging case for unilateral subthalamic deep brain stimulation in Parkinson’s disease. Experimental neurology. 2008;214(1):1–5.

23. Zahodne LB, Okun MS, Foote KD, et al. Cognitive declines one year after unilateral deep brain stimulation surgery in Parkinson’s disease: a controlled study using reliable change. The Clinical Neuropsychologist. 2009;23(3):385–405.

24. Olson JW, Nakhmani A, Irwin ZT, et al. Cortical and subthalamic nucleus spectral changes during limb movements in Parkinson’s disease patients with and without dystonia. Movement Disorders. 2022;37(8):1683–1692.

25. Niccolai L, Aita SL, Walker HC, et al. Correlates of deep brain stimulation consensus conference decision to treat primary dystonia. Clinical Neurology and Neurosurgery. 2021;207:106747.

26. Howell S, Tabibian BE, Mooney JH, et al. Survey of practice preferences in deep brain stimulation surgery in the United States. Interdisciplinary Neurosurgery. 2022;28:101463.

27. Mitchell KT, Larson P, Starr PA, et al. Benefits and risks of unilateral and bilateral ventral intermediate nucleus deep brain stimulation for axial essential tremor symptoms. Parkinsonism & Related Disorders. 2019;60:126–132.

28. Cernera S, Eisinger RS, Wong JK, et al. Long-term Parkinson’s disease quality of life after staged DBS: STN vs GPi and first vs second lead. NPJ Parkinson’s disease. 2020;6(1):1–10.

29. Hewitt KC, Block C, Bellone JA, et al. Diverse experiences and approaches to tele neuropsychology: Commentary and reflections over the past year of COVID-19. The Clinical neuropsychologist. 2022;36(4):790–805.

30. Parsons MW, Gardner MM, Sherman JC, et al. Feasibility and acceptance of direct-to-home tele-neuropsychology services during the COVID-19 pandemic. Journal of the International Neuropsychological Society. 2022;28(2):210–215.

31. Jurica PJ, Leitten CL, Mattis S. Dementia rating Scale-2: DRS-2: professional manual. Psychological Assessment Resources; 2001.

32. Strauss E, Sherman EM, Spreen O. A compendium of neuropsychological tests: Administration, norms, and commentary. American chemical society; 2006.

33. Schmidt M. Rey auditory verbal learning test: A handbook. Vol 17: Western Psychological Services Los Angeles, CA; 1996.

34. Delis DC, Kaplan E, Kramer JH. Delis-Kaplan executive function system. 2001.

35. Bates D, Mächler M, Bolker B, Walker S. Fitting linear mixed-effects models using lme4. arXiv preprint arXiv:14065823. 2014.

36. Rosenthal LS, Salnikova YA, Pontone GM, et al. Changes in verbal fluency in Parkinson’s disease. Movement Disorders Clinical Practice. 2017;4(1):84–89.

37. Højlund A, Petersen MV, Sridharan KS, Østergaard K. Worsening of verbal fluency after deep brain stimulation in Parkinson’s disease: A focused review. Computational and Structural Biotechnology Journal. 2017;15:68–74.

38. Parsons TD, Rogers SA, Braaten AJ, Woods SP, Tröster AI. Cognitive sequelae of subthalamic nucleus deep brain stimulation in Parkinson’s disease: a meta-analysis. The Lancet Neurology. 2006;5(7):578–588.

39. Merola A, Romagnolo A, Krishna V, et al. Current directions in deep brain stimulation for Parkinson’s disease—directing current to maximize clinical benefit. Neurology and therapy. 2020;9(1):25–41.

40. Schnitzler A, Mir P, Brodsky MA, et al. Directional deep brain stimulation for Parkinson’s disease: Results of an international crossover study with randomized, double-blind primary endpoint. Neuromodulation: Technology at the Neural Interface. 2022;25(6):817–828.

41. John KD, Wylie SA, Dawant BM, et al. Deep brain stimulation effects on verbal fluency dissociated by target and active contact location. Annals of Clinical and Translational Neurology. 2021;8(3):613–622.

42. Lin Z, Zhang C, Li D, Sun B. Lateralized effects of deep brain stimulation in Parkinson’s disease: evidence and controversies. npj Parkinson’s Disease. 2021;7(1):1–8.

43. Rohringer CR, Sewell IJ, Gandhi S, et al. Cognitive effects of unilateral thalamotomy for tremor: a meta-analysis. Brain Communications. 2022;4(6):fcac287.

44. Tröster AI, Woods SP, Fields JA. Verbal fluency declines after pallidotomy: an interaction between task and lesion laterality. Applied Neuropsychology. 2003;10(2):69–75.

45. Nijhawan SR, Banks SJ, Aziz TZ, et al. Changes in cognition and health-related quality of life with unilateral thalamotomy for Parkinsonian tremor. Journal of Clinical Neuroscience. 2009;16(1):44–50.

46. Odekerken VJ, Boel JA, Geurtsen GJ, et al. Neuropsychological outcome after deep brain stimulation for Parkinson disease. Neurology. 2015;84(13):1355–1361.

47. Witt K, Daniels C, Reiff J, et al. Neuropsychological and psychiatric changes after deep brain stimulation for Parkinson’s disease: a randomised, multicentre study. The Lancet Neurology. 2008;7(7):605–614.

48. Harati A, Müller T. Neuropsychological effects of deep brain stimulation for Parkinson’s disease. Surgical Neurology International. 2013;4(Suppl 6):S443.

49. Lipski WJ, Alhourani A, Pirnia T, et al. Subthalamic nucleus neurons differentially encode early and late aspects of speech production. Journal of Neuroscience. 2018;38(24):5620–5631.

50. Jorge A, Lipski WJ, Wang D, Crammond DJ, Turner RS, Richardson RM. Hyperdirect connectivity of opercular speech network to the subthalamic nucleus. Cell reports. 2022;38(10):110477.

51. Haynes WI, Haber SN. The organization of prefrontal-subthalamic inputs in primates provides an anatomical substrate for both functional specificity and integration: implications for Basal Ganglia models and deep brain stimulation. Journal of Neuroscience. 2013;33(11):4804–4814.

52. Vonk JM, Rizvi B, Lao PJ, et al. Letter and category fluency performance correlates with distinct patterns of cortical thickness in older adults. Cerebral Cortex. 2019;29(6):2694–2700.

53. Costafreda SG, Fu CH, Lee L, Everitt B, Brammer MJ, David AS. A systematic review and quantitative appraisal of fMRI studies of verbal fluency: role of the left inferior frontal gyrus. Human brain mapping. 2006;27(10):799–810.

54. Askari A, Greif TR, Lam J, Maher AC, Persad CC, Patil PG. Decline of verbal fluency with lateral superior frontal gyrus penetration in subthalamic nucleus deep brain stimulation for Parkinson disease. Journal of Neurosurgery. 2022;1(aop):1–6.

55. Greif TR, Askari A, Maher AC, Patil PG, Persad C. Anterior lead location predicts verbal fluency decline following STN-DBS in Parkinson’s disease. Parkinsonism & Related Disorders. 2021;92:36–40.

56. Temel Y, Blokland A, Steinbusch HW, Visser-Vandewalle V. The functional role of the subthalamic nucleus in cognitive and limbic circuits. Progress in neurobiology. 2005;76(6):393–413.

57. Reich MM, Hsu J, Ferguson M, et al. A brain network for deep brain stimulation induced cognitive decline in Parkinson’s disease. Brain. 2022;145(4):1410–1421.

